# Identifying Incarceration Status in the Electronic Health Record Using Natural Language Processing in Emergency Department Settings

**DOI:** 10.1101/2023.10.11.23296772

**Authors:** Thomas Huang, Vimig Socrates, Aidan Gilson, Conrad Safranek, Ling Chi, Emily A. Wang, Lisa B. Puglisi, Cynthia Brandt, R. Andrew Taylor, Karen Wang

**Affiliations:** Department of Emergency Medicine, Yale University School of Medicine; Section for Biomedical Informatics and Data Science, Yale University School of Medicine; Program of Computational Biology and Bioinformatics, Yale University; SEICHE Center for Health and Justice, Yale School of Medicine; Department of Medicine, Yale University School of Medicine; Equity Research and Innovation Center, Yale University School of Medicine

**Keywords:** Electronic Health Record, Incarceration, Machine Learning, Natural Language Processing, Justice Involvement

## Abstract

**Background:** Incarceration is a highly prevalent social determinant of health associated with high rates of morbidity and mortality and racialized health inequities. Despite this, incarceration status is largely invisible to health services research due to poor electronic health record capture within clinical settings. Our primary objective is to develop and assess natural language processing (NLP) techniques for identifying incarceration status from clinical notes to improve clinical sciences and delivery of care for millions of individuals impacted by incarceration.

**Methods:** We annotated 1,000 unstructured clinical notes randomly selected from the emergency department for incarceration history. Of these annotated notes, 80% were used to train the Longformer-based and RoBERTa NLP models. The remaining 20% served as the test set. Model performance was evaluated using accuracy, sensitivity, specificity, precision, F1 score and Shapley values.

**Results:** Of annotated notes, 55.9% contained evidence for incarceration history by manual annotation. ICD-10 code identification demonstrated accuracy of 46.1%, sensitivity of 4.8%, specificity of 99.1%, precision of 87.1%, and F1 score of 0.09. RoBERTa NLP demonstrated an accuracy of 77.0%, sensitivity of 78.6%, specificity of 73.3%, precision of 80.0%, and F1 score of 0.79. Longformer NLP demonstrated an accuracy of 91.5%, sensitivity of 94.6%, specificity of 87.5%, precision of 90.6%, and F1 score of 0.93.

**Conclusion:** The Longformer-based NLP model was effective in identifying patients’ exposure to incarceration and has potential to help address health disparities by enabling use of electronic health records to study quality of care for this patient population and identify potential areas for improvement.

## INTRODUCTION

Perhaps one of the most underappreciated but highly prevalent social determinants of health is being exposed to incarceration. The United States has one of the highest incarceration rates globally, with over 7 million admissions to jails annually and over 1.2 million in prison as of year-end 2022. ^1–3^ Disproportionately high incarceration rates are observed among racially minoritized individuals, as well as those of low socioeconomic status. Incarcerated individuals have higher rates of communicable and noncommunicable diseases, in addition to mental health and substance use disorders compared with those never incarcerated. ^4,5^ It is estimated that 40 percent of these individuals receive their diagnoses while incarcerated, where there is a constitutional guarantee to health care, but where the acquisition of self-management skills for chronic diseases is hindered by the restrictive and punitive nature of the penal system. ^6^

Upon release, these individuals continue to encounter barriers to care, including limited access to housing, employment, and primary care services. ^7,8^ Compounding these issues, inadequate coordination of care transitions between correctional facilities and community health systems contributes to an elevated risk of death, hospitalization, and deteriorating health outcomes post-release. ^9^ Past work indicates that people with histories of incarceration face significant barriers to accessing consistent and high-quality care, including under-insurance and discrimination within the healthcare system. ^10–12^

These underlying structural factors and social needs drive an important association between increased frequency of acute care utilization and recent or impending incarceration. The frequency of Emergency Department (ED) utilization and frequency of jail encounters per year have been shown to be associated, with those with super-frequent ED use (defined as 18+ visits/year) having 12.3 times the odds of being subsequently incarcerated. In addition, those who were incarcerated saw a significantly increased likelihood of visiting the ED within 30 days prior to incarceration or 30 days following jail exit. ^13^ These interactions with the health system serve as opportunities for health system level interventions to address this social risk, such as engagement in interventions to prevent incarceration (initiation of medications for opioid use disorder, violence intervention programs) or prevent poor outcomes after release (engagement into primary care programs), though screening directly can be stigmatizing. ^14–17^ Additionally, systematically implementing broader health systems level interventions, such as medical legal partnerships, and quality of care analyses necessitate an ability to identify those with a history of incarceration within health system information systems, but currently there is no reliable way to do this.

The electronic health record (EHR) holds promise as a research tool for understanding the drivers of poor health among individuals with a history of incarceration, given the large sample sizes, generalizability to a wide range of patient populations, low expense, and relatively fewer resources needed to conduct studies. ^18^ However, EHRs currently are not designed to systematically measure incarceration exposure. Providers do not receive training in how to ask about or consistently document incarceration history into patients’ social history, leading to current limitations in the documentation of incarceration history in standardized or structured formats. ^19^

Natural language processing (NLP) has the potential to extract valuable information from unstructured data in the EHR, such as in provider notes. NLP techniques, such as named entity recognition, relation extraction, and text classification, can identify relevant information and classify clinical notes according to specific criteria. So far, studies that examine the EHR’s ability to accurately capture data regarding incarceration exposure are limited but demonstrate the potential of this approach. One previous study assessed the identification of incarceration history using an NLP tool, YTEX, on a dataset created through linkage of Veterans’ Health Affairs (VHA) EHR, the Department of Correction (DOC) data, and Centers of Medicare and Medicaid Services (CMS) data. While findings were promising for NLP as an effective means of identification of incarceration history, the study was limited to only VHA EHR which is not generalizable to other health system EHRs. In addition, the YTEX NLP tool is an example of a rule-based NLP in comparison to deep learning techniques for NLP that are able to handle the variability and diversity of human language better in settings utilizing unstructured data, such as clinician notes from the ED. ^18^ Boch et al. proposed a BERT-based model that examined overall parental justice involvement among the pediatric population, demonstrating the utility of NLP in the identification and exploration of justice involvement. ^20^ However, there currently is no tool which identifies an individual’s own history of incarceration and timing of the event based on unstructured clinical encounter notes.

The primary objective of this investigation was to develop an accurate NLP model, using stateof-the-art methods, to reliably and accurately identify incarceration history from unstructured clinical notes in the EHR. We also aimed to create a large database of annotated unstructured clinical notes to serve as a reliable dataset for current and future benchmarking. By pursuing these goals, our investigation will contribute to a better understanding of the utility of NLP techniques for identifying incarceration history in the EHR context, paving the way for improved research on the health of individuals with a history of incarceration and the development of targeted interventions to address their unique health needs.

## METHODS

### Study Population and Setting

The study population consisted of a set of adult patients (*≥*18 years of age) who presented to the emergency department (ED) between June 2013 and August 2021 and had an ED note containing at least one of the following incarceration-related terms: “incarceration,” “jail”, “handcuffs”, “prison”, “incarcerated”, “felony”, “probation”, “parole”, “convict”, “inmate”, “imprisoned”. These terms were defined and selected after a literature review and consultation with expert opinions (LP, EW, KW, RAT). The study was completed across ten EDs within a regional healthcare network in the northeastern United States, covering a geographic area of approximately 650 square miles, and closely resembling the overall national population. ^21^ The study followed the STROBE reporting guidelines for observational studies and was approved by the institutional review board, which waived the need for informed consent (HIC# 1602017249).

### Data Collection and Processing

From an initial set of 81,140 total clinical notes that had at least one of the prespecified key-words, we randomly sampled 1000 notes for annotation, which came from 849 unique patients across 989 visits. The size of this random sample of clinical notes was selected to ensure representation of the diverse presentations and encounter types that a patient with incarceration history could present to the ED with. To ensure model robustness to note type, a total of 25 different note types were selected, the majority of which were ED Provider Notes, Progress Notes, and ED Psych Eval Notes. A full list is in Appendix A. All text was sampled from the system-wide electronic health record (Epic, Verona, WI) using a centralized data warehouse (Helix).

### Defining History of Incarceration

The broad definition of incarceration as the state of being confined in prison or imprisonment, was further stratified into more specific statuses of previous history of incarceration, recent incarceration, and current incarceration. Similar to the process for identifying initial incarceration-related terms, related terms were chosen after an extensive literature review and consultation with expert opinions (LP, EW, KW, RAT). We stratified temporal relationship to incarceration because there are different health risks associated with each. As an example, transition into and out of correctional facilities is disruptive and traumatic and can have differential effects on health. ^22^ Additionally, there are likely different health system level interventions that are feasible to improve care for currently and formerly incarcerated individuals due to the role of departments of corrections in managing health care.

### Document Annotation

The process began with the assembly of a set of provider notes, capturing various clinical encounters related to incarceration and justice involvement. Using the definitions in section “Defining History of Incarceration”, senior authors (KW, AT) defined an initial set of annotation guidelines to determine incarceration status as at least one of three categories: Prior, Current, and Recent incarceration (Table 1). Our team of annotators (TH, CS, LC) led by AT, underwent thorough training on the annotation guidelines. Our annotation process then followed an iterative approach, updating guidelines while classifying an initial set of 50 notes, utilizing Fleiss’ Kappa to evaluate consistency across annotators to ensure a reliable and standardized annotation process throughout the study. Following high reliability between annotators, the remainder of the 1000 notes were randomly distributed among TH, CS, and LC, and the full set was annotated. The task was framed as a classic multilabel text classification task, allowing annotators to select if patient reports had evidence of any of the following: Prior, Current, and Recent Incarceration. If a patient had a history of incarceration and was currently incarcerated, both could be selected.

**Table 1.** Incarceration History Annotation Labels and Corresponding Definitions.

| Label | Definition |
| --- | --- |
| Prior History of Incarceration | Patient has stated history of incarceration (jail or prison), including recent incarceration.<br>Patient has stated current parole status or has a stated history of parole.<br>Patient is stated to be currently living in halfway house specific to incarceration.<br>Note: Label if there is evidence of history of or current parole as well as halfway houses that were explicitly mentioned to serve previously incarcerated individuals |
| Current Incarceration | Patient is stated to be currently in or came from jail or prison. |
| Recent Incarceration | Patient has stated history of being released from jail or prison in the last 6 months.<br>Provider explicitly states recent release from jail or prison. |

Label text classification for “Prior History” was contingent on explicit stated evidence in the note for history of incarceration, or other mentions that could allow for inference that the subject of the note had previously experienced incarceration. This included evidence of history of or current parole as well as halfway houses that were explicitly mentioned to serve previously incarcerated individuals. “Current Incarceration” was coded in instances with confirmed evidence in the text that the subject of the note came directly from a correctional facility. “Recent incarceration” required mention by the author of note stating recent release from a correctional facility or if the language was absent, an explicit mention of date of release as well as date of the note or time since release that fell within 6 months.For annotation we employed Prodigy v1.11.7., a scriptable annotation tool designed to enable data scientists to perform the annotation tasks themselves and facilitating rapid iterative development in NLP projects.

### NLP Development

Once annotations were completed, we initially fine-tuned RoBERTa, a classic BERT-based model, to predict incarceration status in ED notes using Huggingface transformers v4.20.1. However, upon initial inspection, we found that the majority of documents (68.2%) were too long to fit in the context window of classic BERT models (512 tokens, 400 words), reducing performance (shown in Appendix B). Therefore, we utilized an advanced BERT-based model, known as Clinical-Longformer. Transformer-based models leverage self-attention to consider context along the full length of the input sequence. While this provides significant performance improvements, memory consumption enlarges quadratically with sequence length, making analysis of longer documents with classic transformer-based, such as BERT, models computationally infeasible. The Clinical-Longformer model uses sparse attention with a sliding context window, along with reduced global attention for key tokens to reduce memory consumption while keeping performance high and increasing context windows. In particular, we take advantage of the benefits of fine-tuning on domain-specific data and fine-tune Clinical-Longformer on our annotated incarceration status dataset. ^23^ We trained both the classic BERT-based model and ClinicalLongformer model on 800 notes (80%) of the data and evaluated performance on 200 (20%) notes. The model was fine-tuned to predict the presence of any of the categories of incarceration status using a multilabel classification layer added to the top of the base model. We used a binary cross-entropy loss function for training. The training process involved 10 iterations over the dataset with a pre-defined batch size of 16, and gradient descent optimization was utilized to minimize the loss function.

In order to measure the ability of the model to identify incarceration status generally, we collapsed the 3 labels of prior history, current, and recent history of incarceration, to represent any indication of incarceration history. For both settings, we report standard evaluation metrics such as precision, recall, and F1-score to quantitatively measure the model’s performance in identifying incarceration status from the provider notes.

## RESULTS

### Dataset

Of the 1000 notes included which were identified as having at least one incarcerated-related term via keyword, only 559 were found to contain evidence that the patient experienced any history of incarceration, including recent incarceration (137 notes), current incarceration (80 notes), and prior history of incarceration (484 notes). Many notes that were included by simple keyword search for incarceration-related terms but not defined as containing evidence for any history of incarceration included instances where family history of incarceration was documented in the note, other forms of justice involvement, incorrect contexts such as “incarcerated hernia”, and many other examples. Utilizing ICD codes (Z65.1 Imprisonment and other incarceration, Z65.2 Problems related to release from prison) as a means of identification, only 27 of the 562 notes annotated to have any history of incarceration were identified resulting in an accuracy of 46.10%, sensitivity of 4.80%, specificity of 99.09%, precision of 87.10%, and F1 of 0.09 Figure 1.

**Figure 1.**
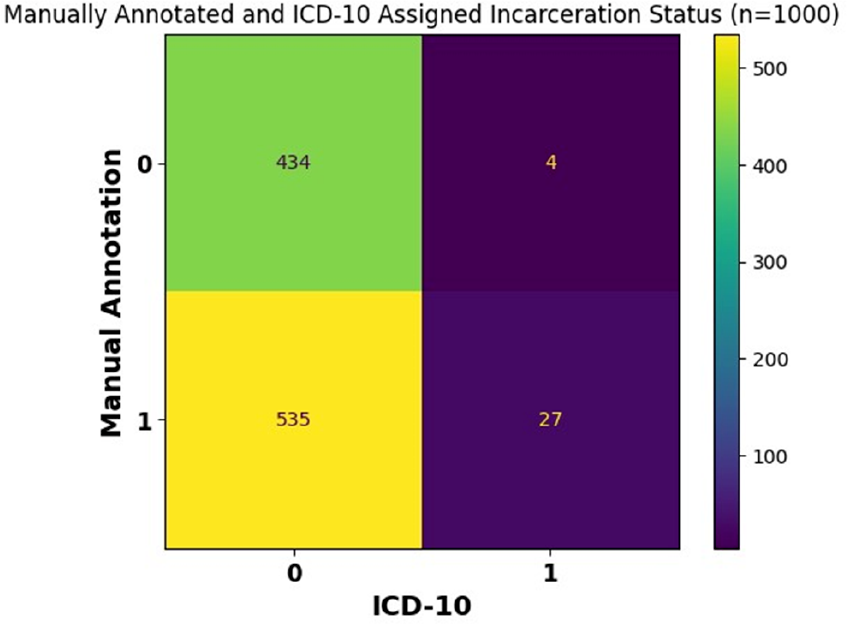
ICD-10 Code vs. Manual Annotation

### Inter-Rater Reliability Performance

To assess the inter-rater reliability, a Fleiss’ Kappa was calculated utilizing overlap of sets of 50 annotated notes between each of the three annotators (TH, CS, LC). The annotators achieved agreement throughout annotating tasks with kappa’s of 0.826 between all annotators. RoBERTa Natural Language Processing To establish a baseline and point of comparison for the ClinicalLongformer model, RoBERTa, another deep learning NLP model, was utilized to identify prior history of incarceration in the test set of 200 manually identified ED encounter notes, recent incarceration, and current incarceration as well as the overall collapsed label of any history of incarceration. For the collapsed label of any history of incarceration, RoBERTa demonstrated an accuracy of 77.0%, sensitivity of 78.6%, specificity of 73.3%, precision of 80.0%, and F1 score of 0.793. Of the total test set of 200 manually annotated notes, there were 22 encounter notes falsely labeled as positive for incarceration history and 24 falsely labeled as negative for incarceration history (Figure 2).

**Figure 2.**
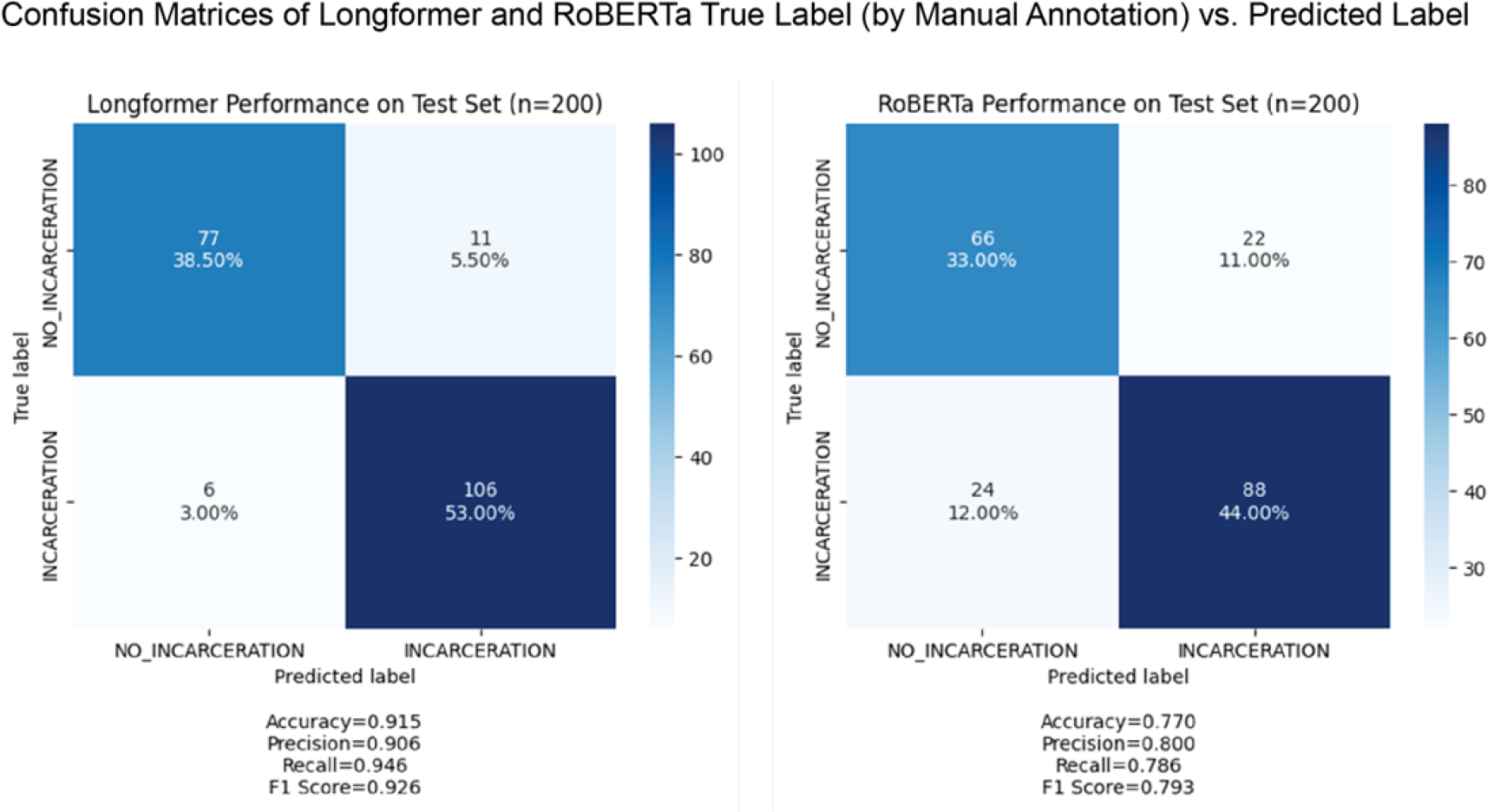
Longformer and RoBERTa Predicted Label vs. True Label by Manual Annotation

As for the more specific temporal labels, RoBERTa demonstrated precision of 78.3%, recall of 75.8%, and F1 score of 0.77 for prior history of incarceration; a precision of 72.4%, recall of 65.6%, and F1-score of 0.689 for recent incarceration, and precision of 56.2%, recall of 56.2%, and F1 score of 0.562 for current incarceration (Appendix B).

### Clinical-Longformer Natural Language Processing

On the same test set of 200 manually annotated notes, the Clinical-Longformer model demonstrated an accuracy of 91.5%, sensitivity of 94.6%, specificity of 87.5%, precision of 90.6%, and F1 score of 0.926 for the identification of any history of incarceration. Figure 2 is two confusion matrices illustrating the performance of both the Clinical-Longformer model (left) and RoBERTa model (right). Of the total 200 individual test encounter notes, 11 notes were falsely identified for a positive incarceration history, and 6 notes were falsely identified as negative for incarceration history.

Similar to the RoBERTa pattern of performance, the Clinical-Longformer model was relatively limited in its ability to identify specific temporal relationships and in distinguishing between prior history of incarceration (precision: 84.9%, recall: 65.3%, F1: 0.738) recent incarceration (precision: 70%, recall: 65.6%, F1: 0.677), and current incarceration (precision: 64.7%, recall: 68.8%, F1: 0.667) (Appendix C).

The behavior of the Clinical-Longformer model was qualitatively assessed through the use of Shapley plots to identify what contextual clues and phrases the model utilizes as signals when identifying incarceration history. These Shapley plots demonstrate tremendous utility for both assessing what elements of an ED encounter notes strongly signal to the model whether a note is positive for incarceration history or negative for incarceration history. These plots are also useful for identifying potential patterns that can cause misidentification, leading to false positives and negatives. This deidentified Shapley plot of an ED encounter note (Figure 3) demonstrates the Clinical-Longformer model correctly identifying incarceration status. Phrases or lines of text that the Clinical-Longformer model often attends to when identifying incarceration history include “in jail”, “in prison”, “released from jail”, “when incarcerated”, “history of being incarcerated.” An interesting pattern of reporting incarceration is when it is used as a time frame, by either the patient or the physician, when discussing illness, medication usage, substance usage, such as “He reports his insulin doses have been incorrect at his prison where he has been incarcerated.”

**Figure 3.**
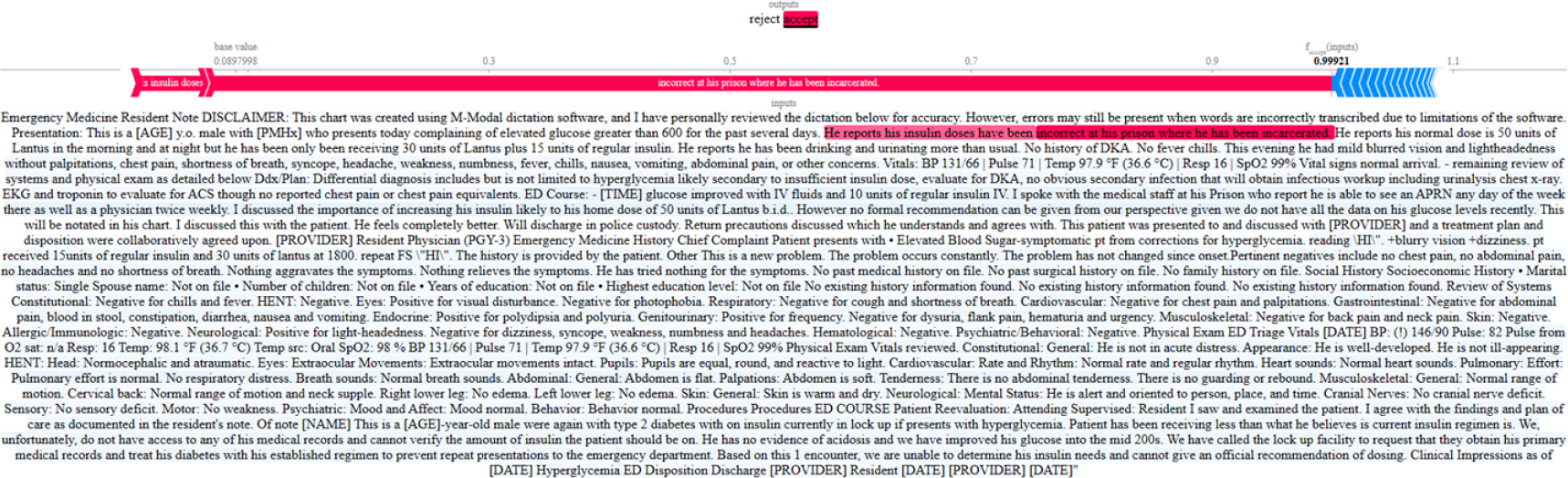
Shapley Visualization of Clinical-Longformer Model for Correct Identification of Any History of Incarceration

Among those 11 notes that were false positives and 6 false negatives for incarceration history, a common trend for the Clinical-Longformer model’s confusion was complex language and phrasing separating current incarceration and instances where the individual was brought in by police or under custody, but not currently incarcerated. While often difficult to even manually annotate, the separation between instances where patients are brought in under a Police Emergency Examination Request (PEER) or from a temporary overnight lock-up is an important difference to distinguish from a patient transported from the carceral system. Phrases such as “She states the patient was kept in the ‘hospital’ part of the jail” confused the model, causing it to be oversensitive in this regard when unable to infer the appropriate context. Other instances of oversensitivity include contextual phrases of “conviction” or “release from court”. This phrasing signals general justice involvement but not necessarily incarceration (Figure 4).

**Figure 4.**
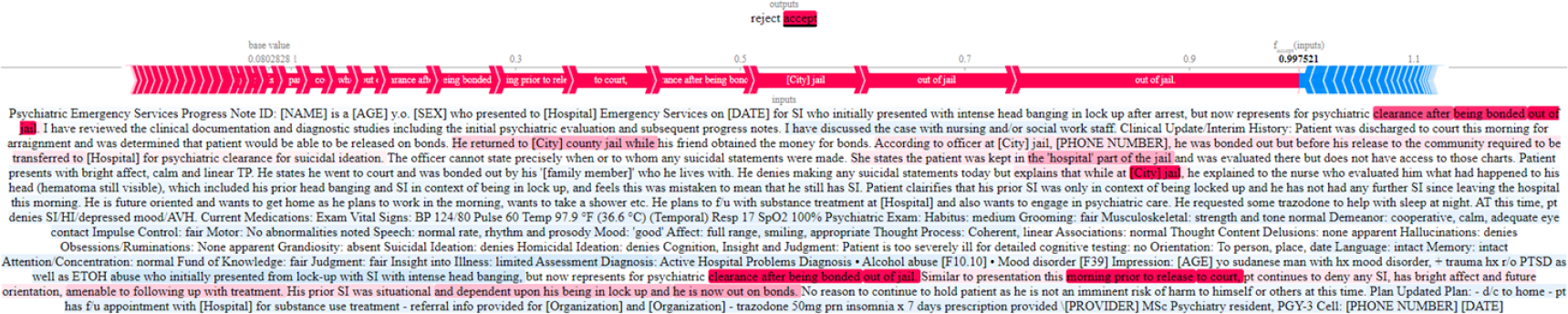
Shapley Visualization of Clinical-Longformer Model for Misidentification of Any History of Incarceration

## DISCUSSION

The criminal justice system, and thus incarceration, is one of the greatest drivers of health inequity that impacts communities across the US. ^24^ Identifying patients with incarceration history within healthcare settings is a key initial step in attending to healthcare inequality and disparity in this underserved patient population. Our Clinical-Longformer Model can reliably and accurately identify incarceration status based on free-form clinician notes in the EHR. This method offers several advantages over other forms of identification such as ICD-10 codes, rule-based NLP techniques, and other NLP techniques like RoBERTa. The NLP algorithm does not rely on providers entering ICD-10 codes which are not used accurately or reliably to measure social determinants of health. The NLP algorithm can capture nuanced information beyond these specific codes through the use of unstructured data when structured data, such as ICD-10 codes and problem lists, often under-report. In addition, the NLP algorithm surpasses simple keyword searches by considering the context and meaning of the text, leading to more accurate identification of incarceration history. ^25^ The Clinical-Longformer model demonstrated superior sensitivity, specificity, precision, and F1 score when compared to the RoBERTa model.

The Clinical-Longformer model developed in this study utilizing deep learning elements offers improvement over previous methods of identification such as the rule-based YTEX model (F1: 0.75), specifically in sensitivity and overall F1 score. ^18^ Additionally, the utilization of a larger training set of 800 unique clinician notes compared to the 228 used in Wang et al., as well as the use of the Clinical-Longformer to improve the attention and analysis over longer notes, likely contributed to the improvement in this NLP model.

Further, our study applies the similar principles utilized by Boch et al. to identify parental criminal justice system involvement in a pediatric population to a more specific and different goal. ^20^ We focused on the identification of incarceration status and history in the subject of the encounter notes while Boch et al. looked at any pediatric exposure to parental justice involvement, including jail, prison, parole, and probation. We focused on narrowing the identification to incarceration history of the patient rather than any justice involvement, further developing our understanding of how NLP can be an asset to healthcare as populations exposed to longterm jail and incarceration histories have unique experiences, health outcomes, and possible interventions through social programs and referrals available to them.

In addition, our Clinical-Longformer model is able to capture and attend to longer documents compared to the BERT model which was used in that study, which is not able to accurately attend to notes over 500 tokens (words) and required significant preprocessing to reduce notes down to snippets containing a total 500 tokens. A study of over 1.6 million ED provider notes, that represented a significant portion (46.2%) of the notes we used for our model, were shown to have an average of 2067 words. ^26^ It is important to note that one token represents 4 characters in English. Thus, the Clinical-Longformer is able to attend well to lengthy ED provider notes and other forms of unstructured data without extensive preprocessing and possible loss of important contextual information that was necessary for the original BERT-based model. Although the metrics of our Clinical-Longformer model are on par with the previous work of Boch et al, the granulation of incarceration history as well as identifying incarceration history specific to the subject of the clinical encounter note can distinguish and help increase the specificity of possible utility for research purposes and possible interventions.

The use of Clinical-Longformer allows for the rapid identification of documented incarceration exposure in the EHR. This information can contribute to a more comprehensive understanding of a patient’s social determinants of health and improve access to real-time referrals to social programs aimed at enhancing healthcare outcomes and finding alternative means of rehabilitation. It can also be used to help guide future research on the potential impact of incarceration on various health outcomes.

The identification of individuals who have had contact with the prison system is the first step in understanding and mitigating disparities in health outcomes for this population. Through the development of models that can help with incarceration history, steps towards improving the quality of healthcare for previously incarcerated patients can be taken as well as addressing existing disparities. ^27^ Previous research has been limited by the difficulty of correctly identifying this population. The use of NLP as a rapid and reliable mechanism to achieve this critical step opens the possibility of future research studies targeting issues such as the disproportionate mortality rate for those diagnosed with cancer during incarceration, or the elevated cardiovascular-related morbidity and mortality for those who have been exposed to the prison system, and providing an opportunity to study quality of care delivery.

The utility of correctly identifying those who have been incarcerated extends beyond research or academic interests. While currently incarcerated patients may be most easily identified in a clinical encounter, those with recent or past history of incarceration often go unidentified, as demonstrated by the poor sensitivity of current implemented systems of identification in ICD-19 codes. Such individuals could benefit by being connected to programs such as the Jail Diversion Task Force, which can help prevent incarceration or re-incarceration and offers rehabilitation to those who would benefit. The Transitions Clinic Network is another evidencebased program available in certain states that acts as a community-based primary care clinic for those returning from incarceration. ^28^ The use of NLP to identify our target population can improve the referral to these programs, as well as encourage the development of additional targeted interventions to help patients avoid imprisonment or reduce the impact of imprisonment on their health. However, the possibility of false positives must also be considered. Care should be taken when approaching patients, no matter how well-intentioned a provider may be, to confirm incarceration history in a non-judgmental way, and to qualify why the provider is asking, before offering resources in order to avoid eroding the patient-physician relationship by contributing to stigma.

While the NLP and machine learning approach for identifying incarceration status shows promise, it is essential to acknowledge its limitations. These limitations include data quality issues, variations in clinician note quality, and potential biases inherent in the algorithm. In addition, the standard for measurement of identification by ICD-10 codes, RoBERTa, and Clinical-Longformer is the compiled manual annotation of three different annotators under the consultation and by the definition developed by both literature review and expert opinion (RAT, KW). Although significant effort and steps were taken to ensure the standard of comparison was representative and consistently applicable, only a total of 1,000 ED encounter notes were manually annotated with a good but not perfect measure of inter-rater reliability. This represents the complexities found within the encounter notes and language when interpreting incarceration status and history.

Hesitancy by patients to disclose incarceration history, as well as hesitancy by providers to include this information in their notes, can lead to underreporting of important incarceration information, rendering the NLP unable to correctly identify incarceration history. Such hesitancy by patients in reporting incarceration history should be heavily considered when utilizing models such as our Clinical-Longformer for identifying patients with incarceration history and applying it in clinical settings. Stigma around incarceration history that is pervasive both within the healthcare system and throughout society at large. The possibility of mis-use of this incredibly powerful tool cannot be ignored. Care should be taken to limit access to this information to those who can be entrusted to work in the patient’s best interest. Thus, the Clinical-Longformer model, given its superior sensitivity and relatively poorer specificity compared to previous models, would more appropriately act as a screening tool or “potential” cohort identifier for further investigation of incarceration history rather than an endpoint of status. Manual confirmation following the use of this Clinical-Longformer model would be best to avoid false positives or misplacements of such electronic labels in a patient file.

In addition, our Clinical-Longformer model was trained over only a small subset of possible ED notes taken from a specific region of the US. While the subset and the annotation were meant to represent the different possible presentations of incarceration history in an unstructured setting, it is possible that this model would not attend well and misrepresent incarceration history in other unstructured data settings such as clinician notes outside of the ED. In addition, with each creation of definitions and annotations, these iterations themselves may add to misclassifications and further decrease the external validity of this NLP model. These misclassifications contributed to a slightly lower specificity in our Clinical-Longformer model when compared to previous YTEX model (99.3%) or ICD-10 code identification. Given the complexity of disparity in healthcare, the impact of incarceration, and stigma surrounding incarceration, any marker for incarceration history should be closely scrutinized.

While the NLP cannot overcome perceived and extant biases in the healthcare system that lead to these documentation shortcomings, our hope is that improving the ease of identifying previously incarcerated individuals for health services research and connection with community programs decreases the stigma around discussions about incarceration. Regarding our NLP itself, while it performs well, it is still early in its development. The majority of phrases used to describe incarceration have likely been captured in this model, however there are certainly other contextual words and phrases that insinuate a history of incarceration that may have been missed and would make this model even stronger.

The Clinical-Longformer NLP was limited in its ability to distinguish the temporal relationship of incarceration based on individual unstructured ED notes. Individually, current, recent, and prior history of incarceration labels were relatively poor in identification compared to identification of “any” prior history of incarceration. Temporal relationships not specific to incarceration have been shown to be difficult to extract using current NLP frameworks. This framework, dependent upon using text from unstructured clinical text taken from a specific time frame, structurally limits the ability for the NLP to extract relevant information to establish temporal relationships. However, although this NLP model was not able to distinguish the temporal relationship of incarceration history based on each individual clinician note, it was still able to accurately identify any history of incarceration. The identification of any history of incarceration, however, is still important in its own right regardless of recency as the very exposure to incarceration is correlated with a wide array of adverse health conditions such as greater self-reported chronic conditions, infectious disease, and mortality. ^29^

Our NLP model serves as a proof-of-concept for future projects aimed at using machine learning to utilize the vast amount of information present in EHR to provide targeted interventions and treatment to patients. Further, improving the ability of this NLP model to attend across multiple notes across data available longitudinally can possibly improve the usage of this model in stratifying incarceration history into distinct sub-periods. The Clinical Longformer here was measured against a dataset of 1000 manually annotated notes based on definitions developed thorough literature review and consultation with experts that was iteratively performed to ensure consistent and reliable annotation. Future application could include measuring this NLP model using linked data systems including EHR and DOC systems.

## CONCLUSION

Our NLP model utilizing Clinical-Longformer with a semi-supervised machine learning approach represents both a reliable and accurate method for identifying incarceration status from nonstructured free form clinician notes in an EHR. It presents several advantages over other methods of identification of incarceration history, such as ICD-10 codes, simple keyword searches, including greater sensitivity, specificity.10 Future research can continue to fine-tune this tool, potentially allowing for the differentiation of current versus previous incarceration in order to better target services and interventions offered to these individuals.

## Data Availability

All data produced in the present study are available upon reasonable request to the authors.

## ACKNOWLEDGMENTS

This publication was made possible by the YCCI Doris Duke Charitable Foundation: Fund to Retain Clinical Scientists (FRCS) and the Yale School of Medicine Fellowship for Medical Student Research.

## APPENDIX

### Appendix A

**Table 2.** Note Type Frequency in 1,000 Annotated Notes.

| Note Type | Count |
| --- | --- |
| ED Provider Notes | 462 |
| ED Notes | 120 |
| Progress Notes | 94 |
| Plan of Care | 88 |
| ED Psychiatric Eval Note | 55 |
| Consult Note | 40 |
| H&P | 35 |
| Admission/Intake | 25 |
| Discharge Summary | 17 |
| Discharge Instructions | 17 |
| ED Observation Note | 17 |
| Discharge Summary | 17 |
| Discharge Summary | 17 |
| Discharge Summary | 17 |
| Discharge Summary | 17 |
| Telephone Communication | 7 |
| SPOC-Behavioral Health | 6 |
| Consults | 6 |
| Operative Note | 5 |
| Evaluation | 4 |
| Telephone Encounter | 3 |
| ED Student Provider | 2 |
| CSC Progress Note | 2 |
| Assessment & Plan Note | 1 |
| Referral | 1 |
| Office/Comment Note | 1 |
| Group Session | 1 |
| H&P (View-Only) | 1 |
| Brief Op Note | 1 |

### Appendix B

**Table 3.** RoBERTa Performance Metrics and RoBERTa Multilabel Performance Confusion Matrix.

|  | Current Incarceration | Prior History Incarceration | Recent Incarceration | macro avg | micro avg | samples avg | weighted avg |
| --- | --- | --- | --- | --- | --- | --- | --- |
| precision | 0.5625 | 0.782609 | 0.724138 | 0.689749 | 0.744526 | 0.3825 | 0.744897 |
| recall | 0.5625 | 0.757895 | 0.65625 | 0.658882 | 0.713287 | 0.3825 | 0.713287 |
| f1-score | 0.5625 | 0.770053 | 0.688525 | 0.673693 | 0.728571 | 0.375667 | 0.728586 |
| support | 16 | 95 | 32 | 143 | 143 | 143 | 143 |

### Appendix C

**Table 4.** Longformer Performance Metrics and Longformer Multilabel Performance Confusion Matrix.

|  | Current Incarceration | Prior History Incarceration | Recent Incarceration | macro avg | micro avg | samples avg | weighted avg |
| --- | --- | --- | --- | --- | --- | --- | --- |
| precision | 0.647059 | 0.849315 | 0.7 | 0.732125 | 0.783333 | 0.3475 | 0.793272 |
| recall | 0.6875 | 0.652632 | 0.65625 | 0.665461 | 0.657343 | 0.3525 | 0.657343 |
| f1-score | 0.666667 | 0.738095 | 0.677419 | 0.69406 | 0.714829 | 0.345 | 0.716525 |
| support | 16 | 95 | 32 | 143 | 143 | 143 | 143 |

